# Psychiatric, Neurological, and Somatic Comorbidities in Intermittent Explosive Disorder: a retrospective cohort study of electronic health records

**DOI:** 10.1101/2024.09.12.24313553

**Authors:** Yanli Zhang-James, John Paliakkara, Joshua Schaeffer, Joseph Strayhorn, Stephen V. Faraone

**Author notes:** Corresponding author: Yanli Zhang-James, MD PhD Associate Professor, Department of Psychiatry and Behavioral Sciences, Norton College of Medicine, SUNY Upstate Medical University 3732B NRB, 505 Irving Ave, Syracuse, NY 13210.

## Abstract

**Importance:** Intermittent Explosive Disorder (IED) is an understudied psychiatric condition that presents with repeated episodes of impulsive aggression and poorly regulated emotional control, often resulting in interpersonal and societal consequences. Better understanding of comorbidities will allow for enhanced screening, diagnosis, and treatment of patients.

**Objective:** To investigate prevalence and associations of IED with psychiatric, neurological, and somatic disorders using real-world data

**Design:** Matched cohorts of patients with or without IED diagnosis were identified using data from the TriNetX Research Network (until January 31, 2024). Cox proportional hazard models were used to estimate and compare the probabilities of acquiring other diagnoses using patients’ available medical records.

**Setting:** Analysis of electronic medical records from two patient populations.

**Participants:** 30,357 individuals with IED and equal number of demographically matched individuals without IED from the TriNetX Research.

**Exposure:** IED diagnosis identified through the associated ICD codes.

**Main Outcomes and Measures:** The main outcomes were ICD-10-CM diagnostic categories and root codes for disorders and health conditions in both cohorts. Main measures are total numbers and proportions of patients who had the diagnostic codes, as well as adjusted hazard ratios for IED diagnosis.

**Results:** Although only 0.03% of the total patient population had an IED diagnosis, we found extensive and widespread comorbidities with psychiatric, neurological and somatic conditions. A significant 95.7% of the individuals with IED had another psychiatric diagnosis. All psychiatric sub-categories and 95% of the psychiatric diagnoses were significantly associated with IED, with HRs ranging from 2 to 77. Among neurological conditions, neurodegenerative diseases and epilepsy had the highest HRs, followed by extrapyramidal and movement disorders, cerebral palsy and other paralytic syndromes, and sleep disorders. Notable associations with IED also includes conditions such as obesity, hyperlipidemia, hypertension, and GERD.

**Conclusion and Relevance:** Our findings illuminate the extensive comorbid relationships between IED and psychiatric, neurological, and somatic disorders. This underscores the necessity for an integrated diagnostic and treatment approach that addresses both the psychological and physical health aspects of IED. Additionally, our work highlights the need for more accurate and inclusive diagnosis of IED in patients with mental disorders.

**Key Points:** Focused Question: What is the pattern of comorbidity of IED with other disorders?

Findings: Despite being underdiagnosed in the clinical cohort, IED was found significantly associated with a wide range of psychiatric disorders, many neurological, and some somatic disorders.

Meaning: The widespread comorbidity highlights the critical need for integrated care approaches that consider the multifaceted health challenges faced by individuals with IED.

## Introduction

Aggressive behavior is an enormous societal problem. How should a psychiatric diagnostic system describe it? Irritability, hostility, disruption, and violence are features of much of mental illness. Psychiatric symptoms are often divided into internalizing and externalizing factors, referring to those harmful to self and others, respectively ^1^. Even disorders typically thought of as internalizing, such as anxiety disorders, may contain “irritability” as a criterion -- the nervous system is not created with clear boundaries between “flight” and “fight.” Between options for coding aggression in a diagnostic system, one would be to define a diagnosis of aggression, noted as present whenever problematic aggression is present, with specifiers for severity and distinctions for impulsive versus instrumental nature. This would allow aggression to be searched easily and its prevalence to be determined. Alternatively, aggression could be considered as a psychiatric problem only when it springs from other psychiatric conditions, without a separate code for aggressive behavior.

The diagnosis of Intermittent Explosive Disorder (IED) represents a compromise between these options, emphasizing the presence of clinically significant impulsive aggression, but only when the aggressive behavior is not better “explained” by another condition, such as bipolar disorder, borderline personality disorder, intoxication or withdrawal from substances. It may be diagnosed in the presence of disorders such as attention deficit hyperactivity disorder (ADHD) or oppositional defiant disorder, but only when the aggression is in excess or warrants independent clinical attention. However, such a definition introduces a logical problem. How can diagnoses such as bipolar disorder or borderline personality disorder “explain” aggressive behavior, when some people with those diagnoses are aggressive, and others are not? How much aggression is “usually seen” in other disorders? These problems are left for individual clinicians and researchers to grapple with, and subjective judgment must play a role in assigning the IED diagnosis.

These considerations pose the question: How do clinicians use the diagnosis of IED: is it applied liberally, or seldom? Is it often applied by itself, or with other psychiatric conditions? What is the pattern of comorbidity? Limited studies have examined the comorbidities of IED with psychiatric disorders, such as bipolar disorder ^2,3^, conduct disorder ^4^, antisocial personality disorder, borderline personality disorder ^4,5^, ADHD ^4,6^, and substance use disorders ^7,8^. Some suggested that IED may be associated with the severity of other psychiatric disorders ^5^, and others considered IED as a potentially useful predictor for identifying and preventing other disorders, given its typically earlier onset compared to many co-occurring conditions ^9,10^.

A comprehensive understanding of the psychiatric comorbidities of IED remains lacking. Much less is known about other medical conditions that often co-occur with IED, especially considering the psychosomatic connections increasingly recognized among mental disorders. An association of IED with sleep disorders has been reported, such as worse symptoms in obstructive sleep apnea and poorer sleep quality scores ^11^. Epileptic patients with IED were reported to have more left temporal atrophy or lesions compared to those without IED ^12^. Most of these studies had limited sample sizes. Our study directly addresses how “real world” clinicians are applying the diagnosis, and what comorbidity looks like in clinical practice, by examining a large dataset of medical records. Our hypothesis is that individuals with IED are more likely to have comorbid health problems, especially other psychiatric conditions with overlapping symptomatology, and somatic conditions, such as infections, injuries, and accidents influenced by their behavior. We aim to illuminate the prevalence and patterns of the full diagnostic co-occurrences in people with IED.

## Methods

### Study population

The TriNetX Research Network contains de-identified data from 117.7 million patients across 87 healthcare organizations (January 31, 2024). The study was determined to be exempt by the SUNY Upstate Medical University Institutional Review Board. We identified 33,410 individuals with at least one diagnostic record of IED (International Classification of Diseases, Tenth Revision, Clinical Modification (ICD-10-CM) code F63.81, or ICD-9-CM codes 312.34, 321.35). We removed those who had their first visits before 1978, were missing the year of birth, or from outside of the US. For every remaining individual with IED, we selected a randomly matched patient who never had diagnostic codes for IED to serve in a control cohort. The matching criteria included age and year at the first record, sex, and US census regions. The final data set contains 30,357 individuals with IED and their demographically matched controls without IED.

### Outcome measures

We examined all ICD-10-CM main categories (designated by letters) and the individual sub-category root codes (designated by letters and two-digit numbers). ICD-9-CM codes were reported as the corresponding ICD-10-CM codes using the General Equivalence Mappings between the two code systems developed by the Centers for Medicare & Medicaid Services ^13^. We also examined the grouped main subcategories and some selected disorders within the mental and behavioral disorders (F category) and the neurological disorders (G category).

### Statistical analysis

We report the total numbers and proportions of patients from each cohort who had the corresponding diagnostic code(s). Cox proportional hazard models (CoxPH) were used to assess the differential risks of acquiring these diagnoses in the IED and Non-IED cohorts while considering the different follow-up time periods. We treated IED as a life-time risk, disregarding the timing of IED diagnosis, because patients with IED can often be diagnosed long after their other known comorbidities, such as bipolar disorder, ADHD, anxiety and antisocial disorder. CoxPH models analyze the probabilities of acquiring each comorbid condition during the observation period where medical records were available and compare the hazard ratios (HRs) of the IED and Non-IED patients adjusted by age, sex, race, ethnicity and regions. For people who are eventually diagnosed with a comorbid condition, only the initial diagnosis, i.e., the first diagnostic record, is used in the analysis, including the one made on the first known visits. For example, a patient may come in the first known visit for an injury resulting from physical altercation at school and later be diagnosed as IED. We considered this as a positive comorbidity event on day one. The same patient may have other injuries later, but those will no longer be considered. For people without the comorbidities of the interest, the end of the observation period is their last known visit. Patients are considered censored at the last known visits if the diagnostic codes were never recorded. A total 1776 diagnostic codes, subcategories or categories of codes were examined, and we report their HRs, 95% confidence intervals (CI) and the log-rank test p value. A Bonferroni corrected p value cutoff 2.82E-05 was applied to assess the significance. All statistical analysis was conducted using STATA 18.

## Results

The total number of patients with IED was 33,410, representing 0.03% of the total patients in the TriNetX Research Network. Table 1 provides detailed demographic characteristics of the IED cohort along with a random sample of 433,131 individuals who never had a diagnosis of IED. Patients with IED were on average eight years younger at their first visit and were more likely to be male, white, and non-Hispanic compared with those without IED. They had more health care encounters (excluding psychotherapy related visits) and a longer history of medical records. 30,357 individuals with IED were successfully matched with individuals without IED. The average age at the first visit was 26 ± 17 years for both cohorts (Table 1). The matched characteristics are predominantly male (70%), white (66%), and non-Hispanic or Latino (66%), and from the south and midwest of the USA. For patients with IED, the average age at the diagnosis of IED was 28 ± 17 years old. The IED cohort had an average 6 ± 5.5 years of medical records. The non-IED cohort had an average 3.7 ± 5 years of records. The median number of visits (excluding psychotherapies) was 143 for patients with IED and 40 for those without IED. More people with IED than those without IED were single (36% vs. 31%).

**Table 1.** Demographic characteristics of the pre- and post-matched cohorts.

|  | Unmatched cohorts |  |  | Matched cohorts |  |  |  |
| --- | --- | --- | --- | --- | --- | --- | --- |
|  | Non-IED | IED | Total | Non-IED | IED | Total | Matched |
| **N (%)** | 433131<br>(93%) | 33021<br>(7.1%) | 466152<br>(100%) | 30357<br>(50%) | 30357<br>(50%) | 60714<br>(100%) | x |
| **age_index (mean, SD)** | 38 (24) | 26 (17) | 37 (24) | 26 (17) | 26 (17) | 26 (17) | x |
| **year_index (mean, SD)** | 2015 (6.5) | 2013<br>(6.3) | 2015 (6.5) | 2014<br>(5.7) | 2014<br>(5.7) | 2014 (5.7) | x |
| **Age at 1st IED Diagnosis (mean, SD)** | . (.) | 28 (17) | 28 (17) | . (.) | 28 (17) | 28 (17) |  |
| **duration_record_years (mean, SD)** | 2.4 (4) | 6.4 (5.9) | 2.7 (4.3) | 3.7 (5) | 6 (5.5) | 4.8 (5.4) |  |
| **total_encounters (non-psychotherapy, mean(SD))** | 34 (102) | 153 (274) | 42 (126) | 40 (110) | 143 (257) | 91 (204) |  |
| **Sex (N, %)** |  |  |  |  |  |  |  |
| **F** | 223640<br>(52%) | 8365<br>(25%) | 232005<br>(50%) | 7903<br>(26%) | 7903<br>(26%) | 15806<br>(26%) | x |
| **M** | 198127<br>(46%) | 23484<br>(71%) | 221611<br>(48%) | 21313<br>(70%) | 21313<br>(70%) | 42626<br>(70%) | x |
| **unknown** | 11364<br>(2.6%) | 1172<br>(3.5%) | 12536<br>(2.7%) | 1141<br>(3.8%) | 1141<br>(3.8%) | 2282<br>(3.8%) | x |
| **Race (N, %)** |  |  |  |  |  |  |  |
| **Others** | 138593<br>(32%) | 6886<br>(21%) | 145479<br>(31%) | 6309<br>(21%) | 6309<br>(21%) | 12618<br>(21%) | x |
| **Black or African American** | 55652<br>(13%) | 4885<br>(15%) | 60537<br>(13%) | 4111<br>(14%) | 4111<br>(14%) | 8222<br>(14%) | x |
| **White** | 238886<br>(55%) | 21250<br>(64%) | 260136<br>(56%) | 19937<br>(66%) | 19937<br>(66%) | 39874<br>(66%) | x |
| **Ethnicity (N, %)** |  |  |  |  |  |  |  |
| **Hispanic or Latino** | 41714<br>(9.6%) | 3304<br>(10%) | 45018<br>(9.7%) | 2711<br>(8.9%) | 2711<br>(8.9%) | 5422<br>(8.9%) | x |
| **Not Hispanic or Latino** | 219572<br>(51%) | 21722<br>(66%) | 241294<br>(52%) | 20152<br>(66%) | 20152<br>(66%) | 40304<br>(66%) | x |
| **Unknown** | 171845<br>(40%) | 7995<br>(24%) | 179840<br>(39%) | 7494<br>(25%) | 7494<br>(25%) | 14988<br>(25%) | x |
| **Region (N, %)** |  |  |  |  |  |  |  |
| **Midwest** | 69282<br>(16%) | 10129<br>(31%) | 79411<br>(17%) | 8677<br>(29%) | 8677<br>(29%) | 17354<br>(29%) | x |
| **Northeast** | 105274<br>(24%) | 8272<br>(25%) | 113546<br>(24%) | 7694<br>(25%) | 7694<br>(25%) | 15388<br>(25%) | x |
| **South** | 176882<br>(41%) | 10523<br>(32%) | 187405<br>(40%) | 10115<br>(33%) | 10115<br>(33%) | 20230<br>(33%) | x |
| **Unknown** | 20966<br>(4.8%) | 1046<br>(3.2%) | 22012<br>(4.7%) | 1029<br>(3.4%) | 1029<br>(3.4%) | 2058<br>(3.4%) | x |
| **West** | 60727<br>(14%) | 3051<br>(9.2%) | 63778<br>(14%) | 2842<br>(9.4%) | 2842<br>(9.4%) | 5684<br>(9.4%) | x |
| **Marital (N, %)** |  |  |  |  |  |  |  |
| **Married** | 69928<br>(16%) | 1950<br>(5.9%) | 71878<br>(15%) | 3463<br>(11%) | 1840<br>(6.1%) | 5303<br>(8.7%) |  |
| **Single** | 112749<br>(26%) | 11737<br>(36%) | 124486<br>(27%) | 9522<br>(31%) | 10964<br>(36%) | 20486<br>(34%) |  |
| **Unknown** | 250454<br>(58%) | 19334<br>(59%) | 269788<br>(58%) | 17372<br>(57%) | 17553<br>(58%) | 34925<br>(58%) |  |
| Note: SD, standard deviation |  |  |  |  |  |  |  |

Table 2 shows that most high-level categories of ICD-10-CM were significantly associated with IED. The mental, behavioral and neurodevelopmental disorders had the highest HR, 3.4, (95%CI: 3.3-3.5), followed by diseases of the nervous system (HR 1.9, 95%CI: 1.8-2.0). Notably, 95.7% of the individuals with IED had another psychiatric disorder, whereas only 28.6% of non-IED had one or more psychiatric diagnoses. Table 3 shows that all psychiatric sub-categories and diseases were significantly associated with IED, with HRs ranging from 2 to 77. Table 4 shows that many neurological subcategories and diseases had significant association with IED, with the neurodegenerative diseases (HR 5.0, 95%CI: 4.1-6.1) and epilepsy (HR 4.9, 95%CI 4.3-5.6) having the highest HRs, followed by extrapyramidal and movement disorders, cerebral palsy and other paralytic syndromes and sleep disorders (Table 4). Four subcategories of neurological diseases were found to not be significantly associated with IED.

**Table 2.** Association with main ICD-10-CM categories.

| **ICD-10 Categories** | **IED-N** | **IED-%** | **Ctrl-N** | **Ctrl-%** | **Adjusted HR** | **CI_L** | **CI_H** | **Uncorrected p-value (Bonferroni significant p bolded)** |
| --- | --- | --- | --- | --- | --- | --- | --- | --- |
| **F01-F99:** Mental, Behavioral and Neurodevelopmental disorders **(excluding IED)** | **29054** | 95.7% | **8691** | 28.6% | 3.4 | 3.29 | 3.51 | **0.00E+00** |
| **G00-G99:** Diseases of the nervous system | **16260** | 53.6% | **6161** | 20.3% | 1.9 | 1.82 | 1.96 | **1.25E-265** |
| **V00-Y99:** External causes of morbidity | **10362** | 34.1% | **4766** | 15.7% | 1.7 | 1.63 | 1.78 | **1.36E-126** |
| **D50-D89:** Diseases of the blood and blood-forming organs and certain disorders involving the immune mechanism | **7497** | 24.7% | **2886** | 9.5% | 1.7 | 1.61 | 1.79 | **1.29E-92** |
| **Q00-Q99:** Congenital malformations, deformations, and chromosomal abnormalities | **5680** | 18.7% | **2334** | 7.7% | 1.7 | 1.56 | 1.77 | **2.31E-59** |
| **I00-I99:** Diseases of the circulatory system | **11881** | 39.1% | **5815** | 19.2% | 1.6 | 1.51 | 1.64 | **7.51E-106** |
| **E00-E89:** Endocrine, nutritional and metabolic diseases | **16328** | 53.8% | **7818** | 25.8% | 1.5 | 1.49 | 1.60 | **6.56E-134** |
| **H00-H59:** Diseases of the eye and adnexa | **9638** | 31.7% | **4524** | 14.9% | 1.5 | 1.42 | 1.54 | **1.09E-73** |
| **K00-K95:** Diseases of the digestive system | **16451** | 54.2% | **8701** | 28.7% | 1.5 | 1.42 | 1.52 | **6.16E-115** |
| **S00-T88:** Injury, poisoning, and certain other consequences of external causes | **18876** | 62.2% | **11341** | 37.4% | 1.4 | 1.38 | 1.47 | **3.09E-110** |
| **N00-N99:** Diseases of the genitourinary system | **11476** | 37.8% | **6110** | 20.1% | 1.4 | 1.33 | 1.44 | **1.21E-62** |
| **H60-H95:** Diseases of the ear and mastoid process | **8680** | 28.6% | **4240** | 14.0% | 1.4 | 1.32 | 1.44 | **8.14E-46** |
| **R00-R99:** Symptoms, signs, and abnormal clinical and laboratory findings, not elsewhere classified | **26040** | 85.8% | **17497** | 57.6% | 1.3 | 1.23 | 1.31 | **1.61E-60** |
| **L00-L99:** Diseases of the skin and subcutaneous tissue | **13283** | 43.8% | **7457** | 24.6% | 1.3 | 1.22 | 1.31 | **3.50E-40** |
| **A00-B99:** Certain infectious and parasitic diseases | **11986** | 39.5% | **6456** | 21.3% | 1.3 | 1.22 | 1.31 | **1.14E-35** |
| **M00-M99:** Diseases of the musculoskeletal system and connective tissue | **17028** | 56.1% | **10806** | 35.6% | 1.2 | 1.21 | 1.28 | **2.40E-43** |
| **J00-J99:** Diseases of the respiratory system | **16540** | 54.5% | **10306** | 33.9% | 1.2 | 1.16 | 1.24 | **1.30E-27** |
| **Z00-Z99:** Factors influencing health status and contact with health services | **24400** | 80.4% | **16252** | 53.5% | 1.2 | 1.16 | 1.23 | **8.13E-33** |
| **C00-D49:** Neoplasms | **5181** | 17.1% | **3404** | 11.2% | 1.1 | 1.03 | 1.14 | 2.11E-03 |
| **O00-O9A:** Pregnancy, childbirth, and the puerperium | **1927** | 6.3% | **1260** | 4.2% | 1.1 | 1.04 | 1.24 | 3.31E-03 |
| **U00-U85:** Codes for special purposes (This includes provisional assignments for new diseases of uncertain etiology or emergency use) | **2340** | 7.7% | **1040** | 3.4% | 1.1 | 0.99 | 1.15 | 1.06E-01 |
| **P00-P96:** Certain conditions originating in the perinatal period | **113** | 0.4% | **74** | 0.2% | 1.4 | 0.93 | 2.15 | 1.09E-01 |

**Table 3.**
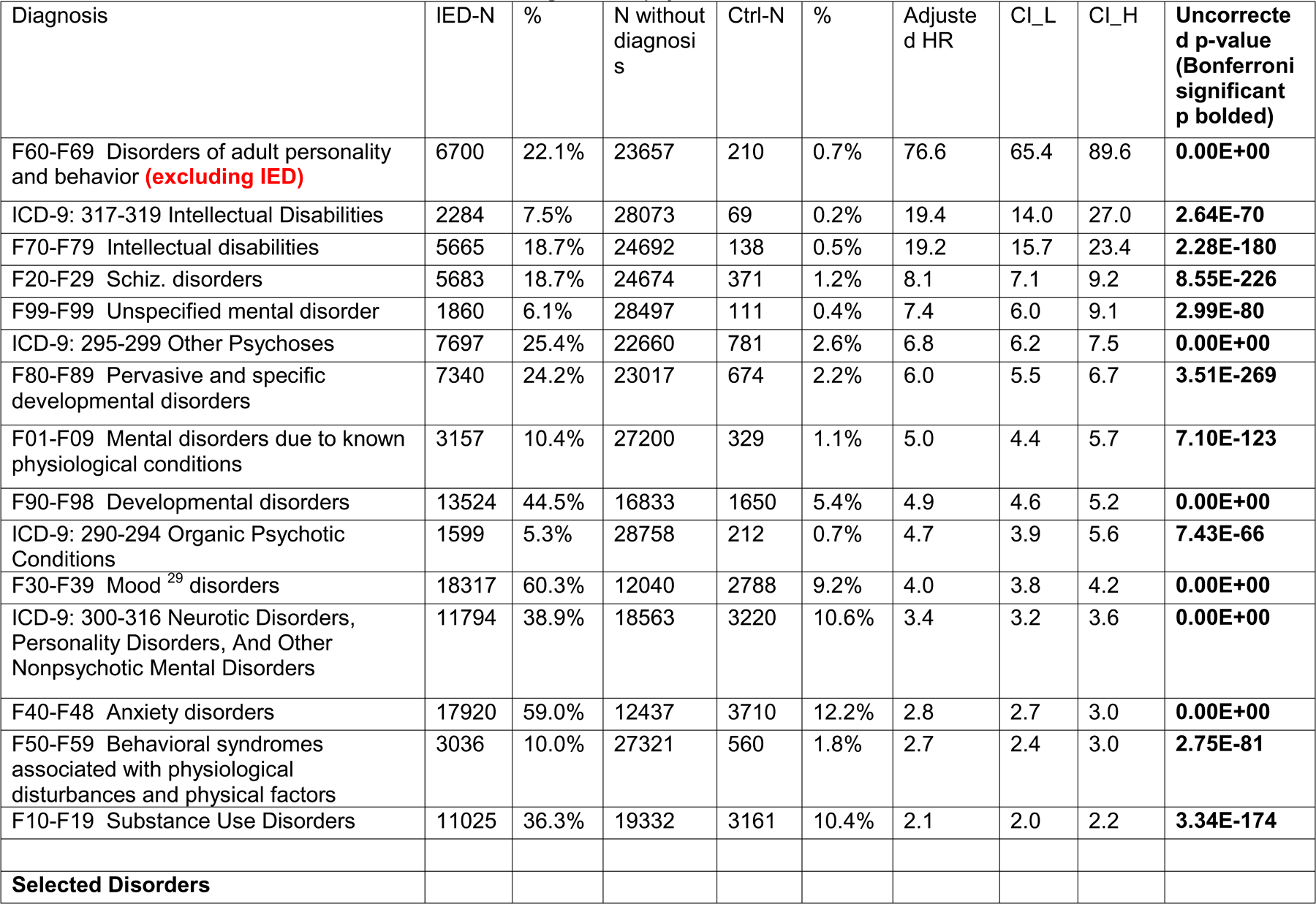
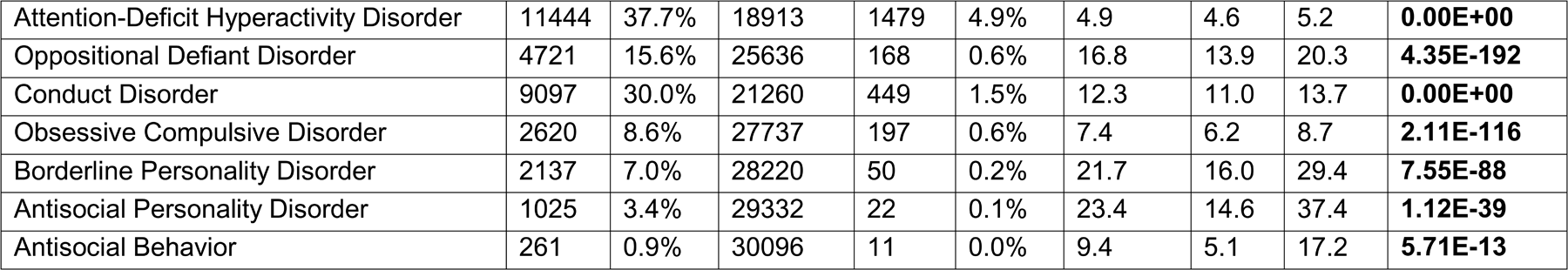
Association with the ICD-10 and 9-CM subcategories for psychiatric diseases and selected disorders.

**Table 4.** Association with main neurological disease subcategories.

| **Diagnosis** | **IED-N** | **%** | **Ctrl-N** | **%** | **Adjusted HR** | **CI_L** | **CI_H** | **Uncorrected p-value (Bonferroni significant p bolded)** |
| --- | --- | --- | --- | --- | --- | --- | --- | --- |
| G30-G32: Other degenerative diseases of the nervous system | 1335 | 4.4% | 162 | 0.5% | 5.0 | 4.1 | 6.1 | **2.22E-54** |
| G40: Epilepsy and recurrent seizures | 4269 | 14.1% | 515 | 1.7% | 4.9 | 4.3 | 5.6 | **7.30E-139** |
| G20-G26: Extrapyramidal and movement disorders | 2575 | 8.5% | 461 | 1.5% | 3.1 | 2.8 | 3.5 | **3.48E-82** |
| G80-G83: Cerebral palsy and other paralytic syndromes | 1591 | 5.2% | 341 | 1.1% | 2.6 | 2.2 | 3.0 | **1.80E-35** |
| G47: Sleep disorders | 8412 | 27.7% | 2156 | 7.1% | 2.2 | 2.1 | 2.3 | **2.68E-168** |
| G70-G73: Diseases of myoneural junction and muscle | 232 | 0.8% | 72 | 0.2% | 2.1 | 1.5 | 2.9 | **1.76E-05** |
| G60-G65: Polyneuropathies and other disorders of the peripheral nervous system | 1954 | 6.4% | 734 | 2.4% | 1.8 | 1.7 | 2.0 | **6.24E-32** |
| G01-G09: Inflammatory diseases of the central nervous system | 1397 | 4.6% | 470 | 1.5% | 1.8 | 1.6 | 2.0 | **2.15E-20** |
| G89-G99: Other disorders of the nervous system | 8030 | 26.5% | 2969 | 9.8% | 1.6 | 1.6 | 1.7 | **2.95E-82** |
| G43: Migraine | 2876 | 9.5% | 1129 | 3.7% | 1.6 | 1.5 | 1.7 | **1.74E-27** |
| G50-G59: Nerve, nerve root and plexus disorders | 1585 | 5.2% | 622 | 2.0% | 1.4 | 1.3 | 1.6 | **5.72E-11** |
| G44: Other headache syndromes | 1291 | 4.3% | 497 | 1.6% | 1.4 | 1.2 | 1.6 | **2.00E-08** |
| G45: Transient cerebral ischemic attacks and related syndromes | 386 | 1.3% | 137 | 0.5% | 1.5 | 1.2 | 1.9 | 1.33E-04 |
| G10-G14: Systemic atrophies primarily affecting the central nervous system | 133 | 0.4% | 36 | 0.1% | 2.4 | 1.5 | 3.8 | 2.55E-04 |
| G46: Vascular syndromes of brain in cerebrovascular diseases | 22 | 0.1% | 5 | 0.0% | 2.4 | 0.9 | 6.6 | 9.58E-02 |
| G35-G37: Demyelinating diseases of the central nervous system | 199 | 0.7% | 122 | 0.4% | 1.2 | 0.9 | 1.6 | 2.62E-01 |

Supplementary Table S1. presents results for all 1723 ICD-10-CM root codes, highlighting that 592 (34.4%) were significantly associated with IED at the Bonferroni corrected p-value threshold, with many psychiatric conditions (F01-F99) showing the highest hazard ratios (HRs).

Figure 1A shows that 92% of the root codes from the F category were significantly associated with IED. Visualization of the significantly associated HRs against their negative log of the uncorrected p-values reveals two clusters (Figure 1B): one with the highest HRs but lower p-values, predominantly including intellectual disabilities, and another with the lowest p-values but low yet still significant HRs, predominantly psychiatric disorders and neurological conditions such as sleep disorders and epilepsy. Figure 1C plots the prevalences of the significantly associated disorders in both the IED and non-IED populations, highlighting the highly prevalent comorbidities, including anxiety, depression, ADHD, and sleep disorders.

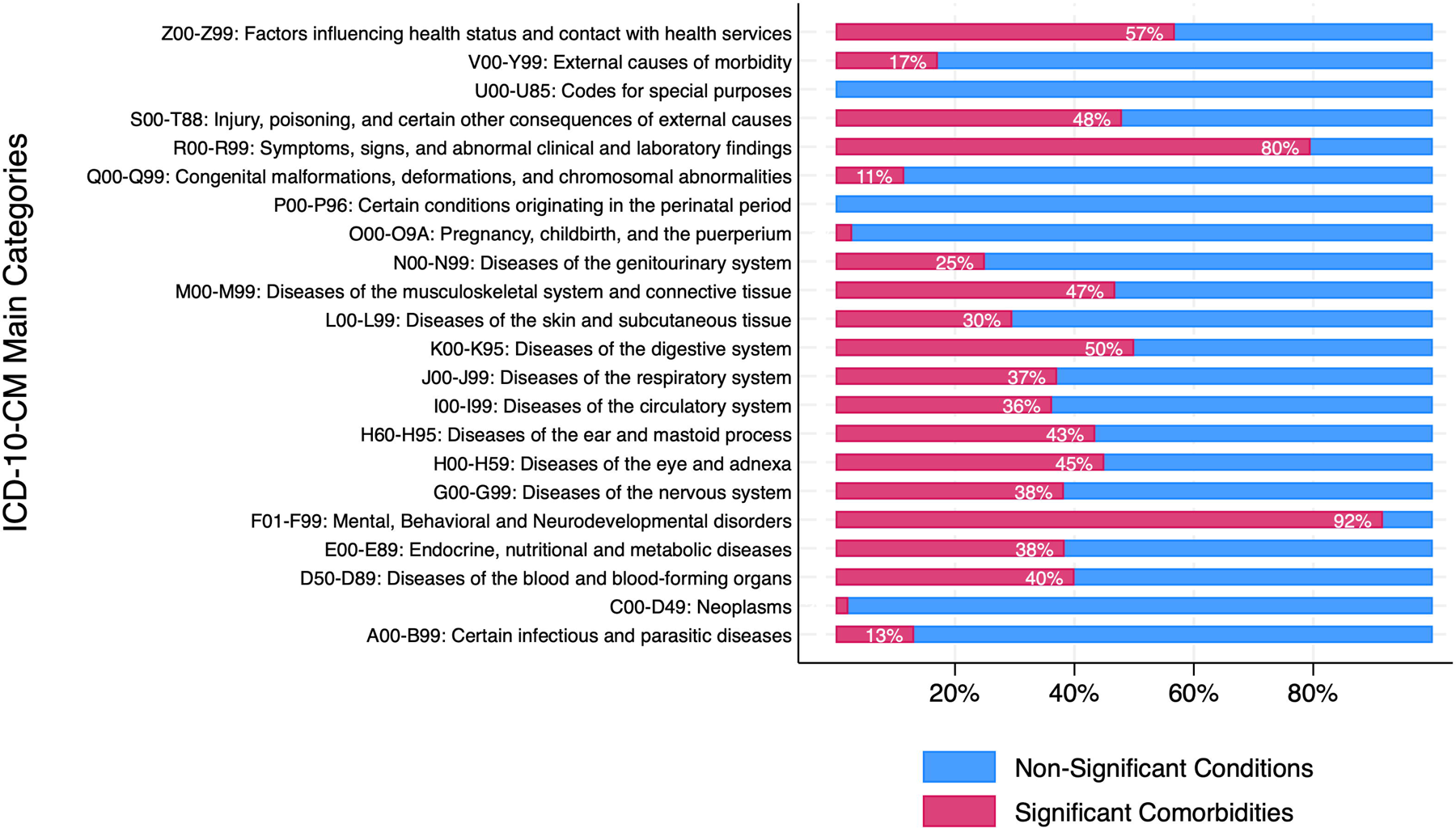
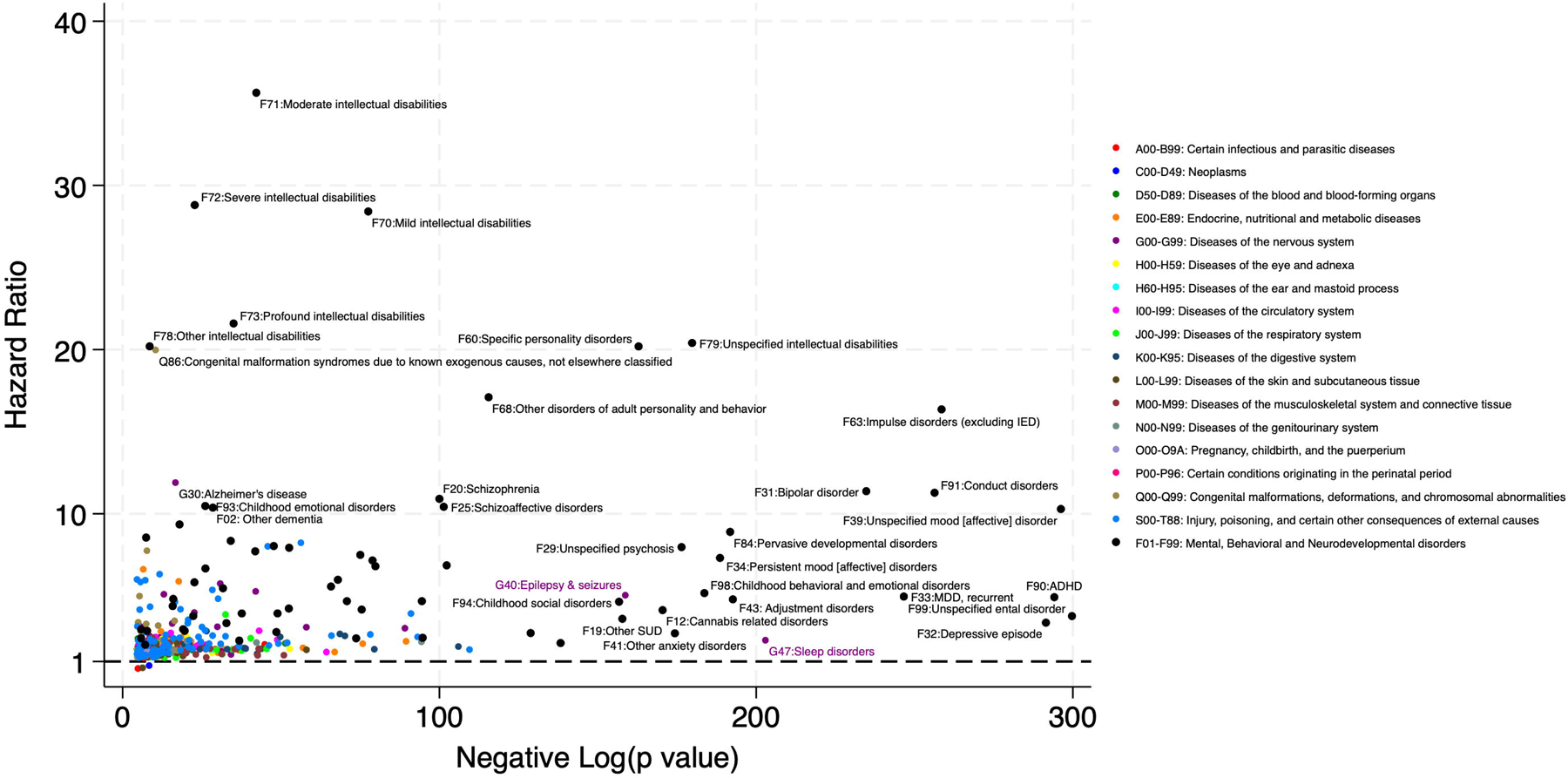
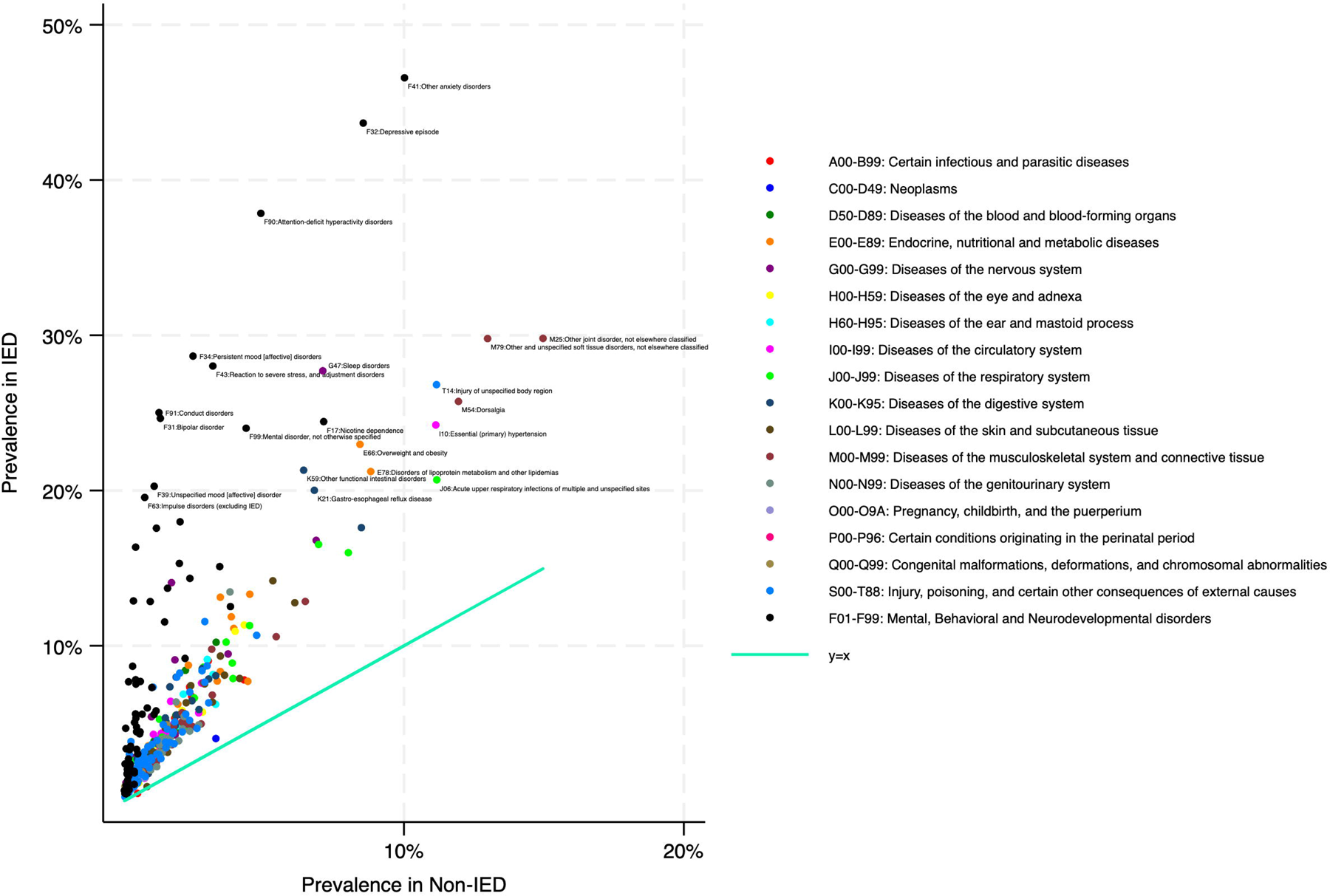

## Discussion

In our medical record data from over 100 million patients, the lifetime prevalence of IED was only 0.03%, in marked contrast to the 7.3% lifetime prevalence reported from the USA National Comorbidity Replication Study ^9^, and even the 0.8% lifetime prevalence reported in a multinational study ^14^. The prevalence of problematic aggression or violence, much of which may be “impulsive” aggression), is surely still higher. Sturmey (2022) noted “Aggression and violence are widespread in humans with around half a million murders occurring per year globally, mostly involving young men killing one another. About one third of women also experience aggression and violence, including sexual violence, often during interpersonal relationships during their lifetime. About half of all children worldwide experience various forms of aggression and violence often from family members and in educational settings” ^15^. A community survey reported that 12% of people disclosed having “trouble controlling their own anger” ^16^. Despite the “better explained by” exclusion in diagnostic criteria, we believe that if clinicians diagnosed IED whenever patients met existing criteria, the prevalence would have been much higher, similar to the rates found in epidemiologic studies. Our data do not provide clues about the reasons for the rarity of the diagnosis in clinical practice.

Although the DSM-V criteria specify that for a diagnosis of IED, the outbursts should not be “better explained” by a “psychotic disorder, antisocial personality disorder, borderline personality disorder,” our sample showed high comorbid with each of these conditions. IED can be diagnosed alongside ADHD when “recurrent aggressive outbursts are in excess of those usually seen… and warrant independent clinical attention.” About 38% of the IED patients in our data had ADHD diagnoses. Although the criteria state that the outbursts should not be attributable “to the physiological effect of a substance,” about 15% had alcohol use diagnoses, compared with about 3.5% of the non-IED group. Although cannabis is thought to be much less highly related to aggression than alcohol (questioned by Rafiei & Kolla ^17^), 15% of IED patients had cannabis related disorders. Although IED is a quintessential “externalizing” disorder, it was strongly comorbid with both anxiety and mood disorders.

Although our analysis did not examine the timing of these other psychiatric diagnoses relative to IED, it is remarkable that every category of psychiatric morbidity was found more frequently among IED patients compared with matched controls (Table 3), and that 92% of all F category root codes were significantly associated with IED (Figure 1). Many of the differences were substantial; HRs ranged from 2 to 77. A striking finding was that IED without psychiatric comorbidity, or “pure” IED was rare: only 4.3% of those with IED did not have another psychiatric diagnosis (Table 2). Of the psychiatric categories, the largest hazard ratios were for intellectual disabilities and personality disorders. Even though the HRs were low, albeit significant, for some disorders, the prevalence of some of these disorders was very high in the IED group (e.g., developmental disorders 45%, mood disorders 60%, anxiety disorder 59%).

The widespread comorbidity with other psychiatric conditions is not surprising, given that outbursts of anger, aggression, irritability, and hostility are present in the criteria or associated features of many psychiatric disorders. One of us found, in a page-by-page count of the DSM-5, 52 such disorders ^18–20^. Given the huge importance of aggression for society, further research might explore whether psychiatric diagnostic systems should include a specifier for rating the severity of aggressive behavior across diagnoses.

The extent of psychiatric comorbidity with IED in our finding also aligns with research on the structure of psychopathology that posits a single “p factor” analogous to the “g factor” of intelligence. Caspi et al. (2014) suggest “Almost all of the variation in the lower-order abilities is accounted for by p” ^1^. They also note the non-specificity of etiological influences on psychopathology, stating that “Indeed, it is more difficult to identify a disorder to which childhood maltreatment is not linked than to identify a disorder to which it is linked with specificity.” Our findings support a similar conclusion about IED, and probably impulsive aggression in general: it is difficult to find a psychiatric problem to which it is not linked.

Many other medical conditions were also found more frequently in IED patients. Notably neurological conditions, including epilepsy, sleep disorders and migraine, were highly prevalent, ranging from 10 to 28%, compared to only 2-7% in those without IED. Interestingly, neurodegenerative disorders, despite being at lower prevalence (4% vs. 0.5%), had some of the highest HRs. This is noteworthy even though DSM criteria for IED specify that the outbursts should not be “attributable to another medical condition (e.g., head trauma, Alzheimer’s disease).” It is possible symptoms of IED predate these health issues. In examining comorbidities (Table S1), those most highly associated with IED tend to be more behavioral in nature, such as falls, burns, poisonings, injuries, and “problems related to lifestyle.” In contrast, conditions not significantly associated with IED tend to be more medical in nature, such as gout, acute appendicitis, kidney stones, multiple sclerosis, lupus, and melanoma in situ.

Notable medical associations, such as obesity, hyperlipidemia, hypertension, and gastroesophageal reflux disease (GERD), have not been reported for IED previously. Also significant were associations with various Z category non-diagnostic codes, indicating psychosocial (e.g., childhood trauma) and socioeconomic hardships, consistent with previous research ^4,14,21,22^. Not uniquely, many psychiatric disorders also share similar psychosocial risks ^23,24^. Obesity, linked to adverse psychosocial factors, may also contribute to other medical issues like GERD ^25–28^. The extensive comorbidity is no doubt complex and of multifactorial nature. Nevertheless, our study is the first in using “real-world” data to document these relationships with IED.

Our results should be interpreted considering several limitations. Most notably, we relied on medical records, which, although allowing us to examine a large sample of IED patients, no doubt, created a sample significantly from one that would have been gathered through structured interviews. Validation of our findings through prospective epidemiologic studies is warranted. Given the very low prevalence of IED in our sample, some patients with IED may have been included in the non-IED cohort, potentially diluting our findings and reducing statistical power. Conversely, those with known IED diagnosis may represent more severe cases, magnifying the associations with comorbid psychiatric and somatic disorders. Future studies with confirmatory IED diagnosis and symptom severity measures are needed to validate our findings.

Despite limitations, our findings shed unique light on how IED is diagnosed in clinical practice, distinct from research settings. They raise provocative hypotheses for clinical practice. First, it would be helpful for clinicians to apply the diagnosis of IED more frequently when warranted. Highlighting the problem of aggression in a separate diagnosis may focus more attention on aggressive behavior and the development of specific treatments. Otherwise, aggressive behavior remains somewhat hidden, tucked away as a feature that may or may not be present in many different disorders. Second, the comorbidity of IED with falls, poisonings, burns, injuries and accidents suggests that the lack of impulse control may bring people to medical attention for various other conditions. The fact that 34% of ICD codes were significantly associated with IED raises the hypothesis that impulsivity may be an underestimated risk factor in health care in general. The ideal treatment for IED may include addressing general “thinking before acting” skills and not just aggressive behavior. Third, the association between IED and ADHD, with both centered on the trait of impulsivity, raises the hypothesis that conscientious and thorough treatment of ADHD from an early age could conceivably reduce the prevalence of IED.

The findings also raise questions regarding the construction of diagnostic systems. Given the “better explained by” exclusions for IED, and given the very high psychiatric comorbidity, it is impossible to know how often IED is overlooked, versus not given due to clinicians’ judgment that aggression is better explained by another diagnosis. Allowing IED to be diagnosed regardless of comorbid psychopathology would make it easier to track the prevalence and recovery rates of problematic aggression and would avoid the illogic of declaring that aggression is “explained by” conditions wherein some people are aggressive, and others are not.

We believe that impulsive aggression, defined with or without exclusion criteria for other psychiatric conditions, and diagnosed liberally or as a last resort, will remain robustly comorbid with a wide variety of psychiatric and other medical conditions. This hypothesis should be confirmed or denied by future research.

## Data Availability

All data produced in the present work are contained in the manuscript and available online at MedRxiv. Patient medical records were only available directly from TriNetX.

## Data Access, Responsibility and Analysis

Yanli Zhang-James had full access to all the data in the study and takes responsibility for the integrity of the data and the accuracy of the data analysis.

## Author Contributions

YZJ – conceptualization, methodology, formal analysis, resources, writing, visualization, interpretation, supervision, project administration; JP – writing, literature research, interpretation and editing; J Schaeffer – writing, literature research, interpretation and editing; J Strayhorn – conceptualization, analysis, writing, interpretation and editing; SVF - conceptualization, methodology, analysis, writing, interpretation, editing and supervision

## Data Sharing Statement

The individual-level electronic health record data in the TriNetX database cannot be shared, as they are the property of TriNetX and can only be accessed through direct contract with the company. Summary data and results are provided in the Tables and Supplementary Tables provided in the manuscript.

## Artificial Intelligence

No AI tools were used in the writing or editing of this manuscript

